# A Pilot Study Observing High Salivary Non-Secretor (se/se) Prevalence in Breast and Ovarian Cancers in Kolkata

**DOI:** 10.1101/2025.09.16.25335850

**Authors:** Adrija Das Biswas, Swarnabindu Banerjee, Prasun Bhattacharya

**Affiliations:** Department of Immunohematology and Blood Transfusion (IHBT) Medical College and Hospital, Kolkata (West Bengal University of Health Science); Department of Medical Oncology Medical College and Hospital, Kolkata; Head of the Department Immunohematology and Blood Transfusion (IHBT) Medical College and Hospital, Kolkata

**Keywords:** FUT2 gene, non-secretor, breast cancer, ovarian cancer

## Abstract

**Background:** Breast and ovarian cancers remain major causes of illness and death in women. Secretor status determined by fucosyltransferase 2 (FUT2) gene controls the presence of ABO antigens in all bodily fluids except CSF, and prior studies suggest non-secretor status may be associated with cancer risk. Data on secretor prevalence in breast and ovarian cancers are limited.

**Methods:** We performed a pilot case–control study at a government medical college in Kolkata between January and June 2024. The study included 74 women: 37 patients with newly diagnosed breast (n = 23) or ovarian (n = 14) cancer and 37 healthy female blood donors. Unstimulated saliva was collected and tested for secretor status using a hemagglutination inhibition assay. Associations were tested with the chi-square test.

**Results:** Among healthy controls, 78% were secretors and 22% were non-secretors. In contrast, 81% of cancer patients were non-secretors and 19% were secretors. By cancer type, non-secretor prevalence was 82.6% in breast cancer and 78.5% in ovarian cancer. The association between non-secretor status and cancer was statistically significant (p < 0.0001).

**Conclusions:** In this pilot study, salivary non-secretor status was much more common in women with breast or ovarian cancer than in healthy controls. The findings support larger, population-level and genetic studies to confirm whether FUT2 non-secretor status could serve as a simple, low-cost marker to identify the population at risk. Limitations include small sample size and absence of FUT2 genotyping.

## INTRODUCTION

With notable geographical variations in incidence, mortality, and access to care, female cancers remain a growing global public health concern. Breast cancer accounts for about 25% of all female cancer occurrences worldwide, with 2.3 million new diagnoses in 2022 alone. Despite being less common ovarian cancer still causes over 50% of all gynecological cancer-related fatalities, primarily because of delayed diagnosis and limited treatment options. These results highlight the critical need for better risk classification and early detection techniques, especially in low- and middle-income nations where resource limitations and healthcare inequities persist.^1^

Aberrant glycosylation, particularly fucosylation, has emerged as a key factor influencing cancer susceptibility by regulating ABO blood group antigen expression in secretions, shaping mucosal immunity, maintaining epithelial integrity, and modulating microbiome composition.^2–5^Alterations in these processes can lead to immune evasion, microbial dysbiosis, chronic inflammation, and disrupted tissue integrity, all of which have been shown to promote tumor initiation and progression.^6–8^

Secretor status, determined by the FUT2 gene on chromosome 19q13.3, dictates the presence or absence of A, B, and H antigens in bodily secretions (Figure 1). Individuals with functional FUT2 alleles (Se/Se or Se/se; secretors) produce these antigens in saliva and mucus, whereas non-secretors (se/se) lack them despite antigen presence on erythrocytes via FUT1^9^. Approximately 20-30% of the healthy population worldwide are non-secretors while 70-80% are secretors.^10–12^

**Figure 1:**
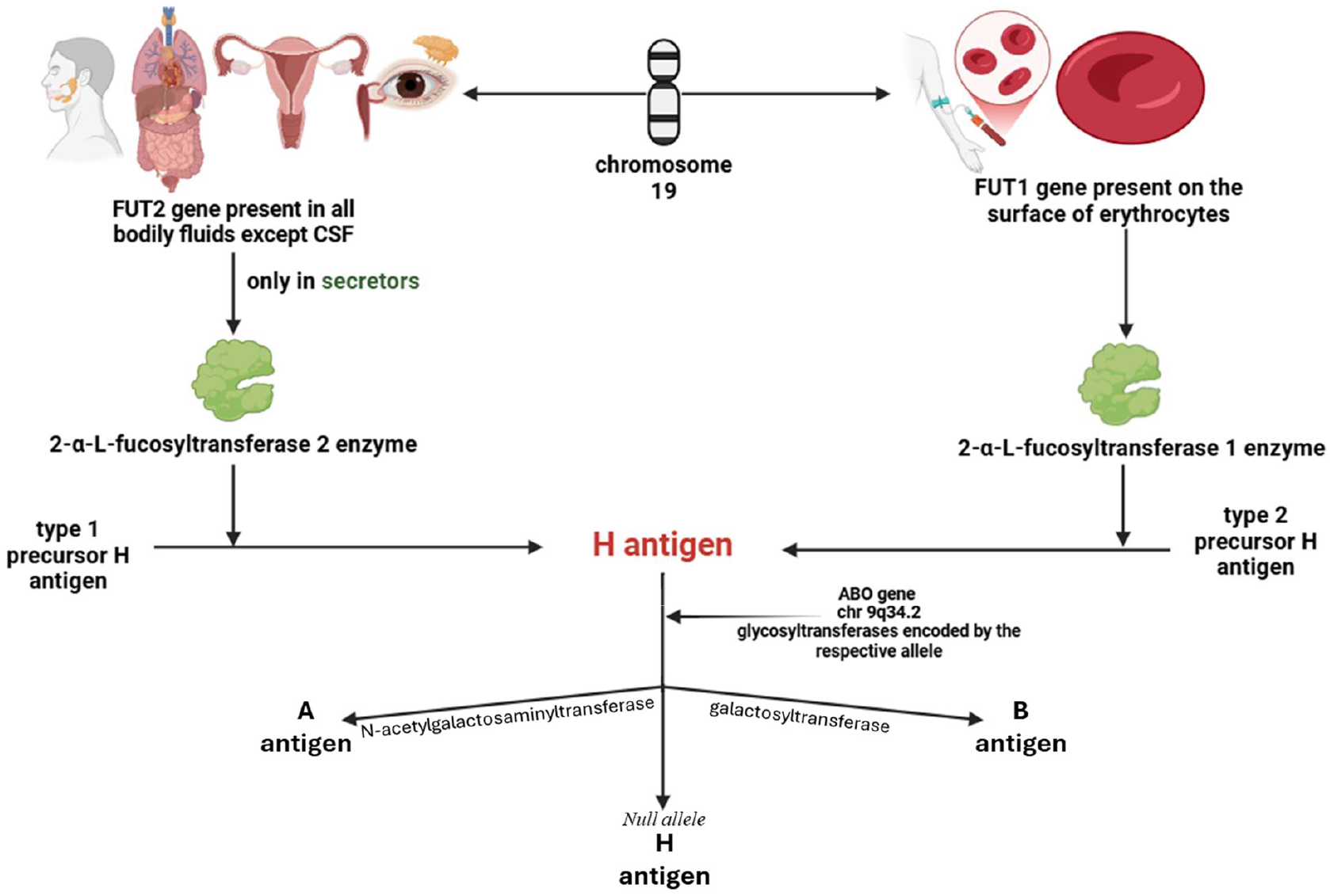
Biosynthesis and expression of ABO blood group antigens regulated by FUT1, FUT2, and ABO genes. FUT1 encodes α-1,2-fucosyltransferase 1, producing H antigen on erythrocytes. FUT2, expressed in secretory epithelial cells, encodes α-1,2-fucosyltransferase 2, forming H antigen in secretions; only secretors express this enzyme. The ABO gene modifies H antigen to A or B antigens, or results in O blood type if null.

Several serological studies have reported a higher prevalence of non-secretors among patients with various cancers, suggesting an association between secretor status and cancer susceptibility.^13–16^ In addition, genetic association studies have explored links between FUT2 variants and cancer risk, further supporting the role of secretor status in tumorigenesis.^17–20^

Despite this growing body of evidence and the substantial global burden of breast and ovarian cancers, data on secretor prevalence in these malignancies remain limited. In addition to providing prevalence estimates, such data could help determine whether secretor status exerts similar effects across multiple cancer types, with important implications for understanding its role in tumor biology and expanding its potential clinical applications.

This pilot study aimed to estimate the serological prevalence of FUT2-mediated non-secretor status in breast and ovarian cancer patients relative to healthy controls. To the best of our knowledge, based on available literature, this is the first study to assess the seroprevalence of non-secretor status in breast and ovarian cancer patients, providing foundational evidence to guide subsequent molecular and population-based studies. Addressing this gap is essential for advancing our understanding of glycosylation-mediated mechanisms in tumorigenesis and exploring the potential of salivary secretor status as a marker to identify the population at risk. By providing a non-invasive and cost-effective approach, saliva-based assessment could facilitate population-level screening, particularly in high-burden, resource-limited settings.

## Materials and Methods

### Study Design and Participants

This case-control study was conducted at the Departments of Immunohematology and Medical Oncology, Medical College and Hospital, Kolkata, between January 2024 and June 2024. Ethical clearance was obtained from the Institutional Ethics Committee (MC/KOL/IEC/NON-SPON/2211/11/2023), and written informed consent was obtained from all participants.

A total of 74 female participants were included in the study (Figure 2):

**Figure 2:**
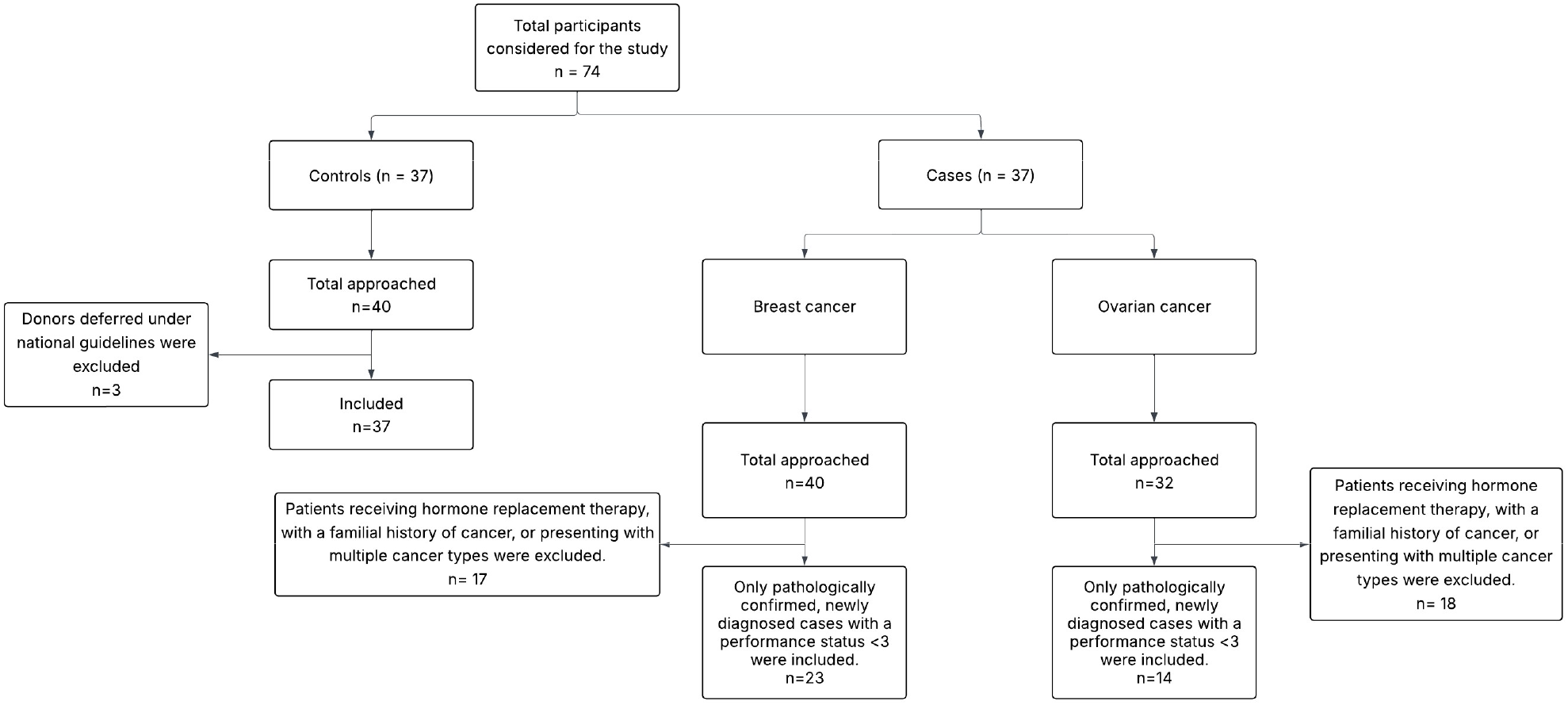
Flow diagram of study participant selection. A total of 74 female participants were considered, including 37 cases (23 breast cancer, 14 ovarian cancer) and 37 healthy controls. The diagram illustrates the number of participants initially approached, excluded, and ultimately included in the study.

- Cases: 37 female patients (23 with breast cancer and 14 with ovarian cancer) were enrolled using purposive sampling.
- Controls: 37 healthy female blood donors were enrolled via simple random sampling from blood donation camps.

### Inclusion and Exclusion Criteria

#### Cases (Breast and Ovarian Cancer)

- Inclusion: Newly diagnosed, pathologically confirmed breast or ovarian cancer; performance status <3.

Ensures that all cases have a definitive diagnosis. By including only newly diagnosed patients, the study avoids survival or treatment-related biases. Patients with a reasonably good functional status are included to reduce confounding due to severe illness or comorbidities

- Exclusion: Patients receiving hormone replacement therapy, with a familial history of cancer, or presenting with multiple cancer types.

HRT can influence hormone-sensitive cancers and individuals with hereditary cancer predisposition may have distinct genetic profile. Presence of multiple malignancies may introduce heterogeneity.

#### Controls

- Exclusion: Donors deferred under national guidelines, as per the Drugs and Cosmetics Act and the National Blood Transfusion Council, Ministry of Health and Family Welfare, Government of India. controls.

### Participant Attrition

40 breast cancer patients and 32 ovarian cancer patients were initially approached. After applying the inclusion and exclusion criteria, 23 breast cancer cases and 14 ovarian cancer cases were included (Figure 2). Similarly, 40 healthy donors were approached for the control group, of whom 3 were excluded due to deferral under national guidelines, resulting in 37 controls.

### Ethics Approval and Consent to Participate

Ethical clearance for this study was obtained from the Institutional Ethics Committee, Medical College and Hospital, Kolkata (MC/KOL/IEC/NON-SPON/2211/11/2023). Written informed consent was obtained from all participants prior to sample collection. All procedures were conducted in accordance with the Declaration of Helsinki and relevant guidelines.

### Sample Collection

Unstimulated saliva samples (≥3 mL) were collected from participants into sterile containers. Participants were instructed to provide saliva, without chewing or ingesting any substances prior to collection. Samples were aliquoted, centrifuged at 3,000 rpm for 10 minutes, and the supernatant preserved in RIA vials. The supernatant was boiled at approximately 100°C for 5– 6 minutes, re-centrifuged, diluted with an equal volume of normal saline, and stored at –20°C until analysis.

### Assessment of Secretor Status

Secretor status was determined using the hemagglutination inhibition assay, following standard operating procedures from the Department of Immunohematology and the 21st edition of the AABB Technical Manual (2023).^21^

#### Principle

Secretors express A, B, or H antigens in saliva. During the inhibition assay, these antigens bind the respective antisera, neutralizing its agglutination potential. When a 5% RBC suspension is added, the absence of agglutination indicates the presence of the secreted antigen.

The principle of the hemagglutination inhibition assay for determining secretor status is illustrated in Figure 3.

**Figure 3:**
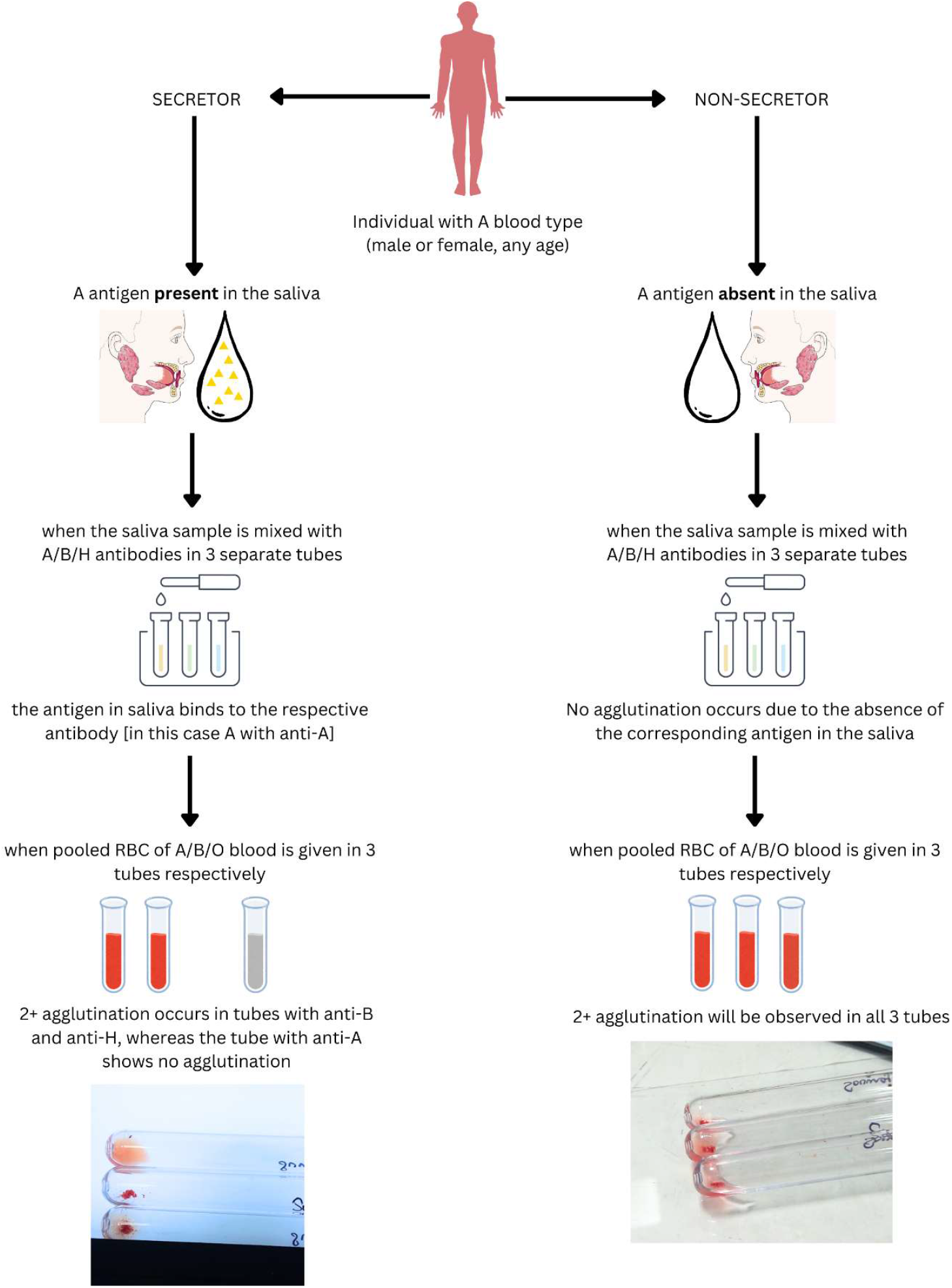
Schematic representation of hemagglutination inhibition assay outcomes in secretor and non-secretor individuals (blood group A). In secretors (left), the A antigen present in saliva binds to anti-A antibody, preventing agglutination. Consequently, no agglutination is observed in the anti-A tube, while agglutination occurs in the anti-B and anti-H tubes. In non-secretors (right), salivary A antigen is absent, leaving anti-A antibodies free to react with red blood cells, resulting in agglutination in all three tubes.

##### I. Serial Dilution

For the serial dilution, three blood group–specific setups (A, B, O) were prepared for each sample. A total of 24 RIA vials were arranged in two parallel rows of 12, representing Mother and Daughter vials. Ten vials in each row were designated for a logarithmic dilution series ranging from 1:2 to 1:1024, with the first vial containing the neat sample and the last vial serving as a blank control. Two-fold serial dilutions were performed by adding 200 µL of monoclonal/polyclonal antibody (A/B/H) to the neat vial, followed by sequential transfer of 100 µL across vials pre-filled with 100 µL of normal saline. The final 100 µL from the 1:1024 dilution was transferred into the blank vial.

##### II. Addition of RBC Suspension

Subsequently, 50 µL of 5% RBC suspension was added to each Mother vial for the respective blood group, and the vials were centrifuged at 1,000 rpm for 1 minute. The first dilution exhibiting 2+ agglutination (zone of equivalence) was identified, and the corresponding Daughter vial was used for the hemagglutination inhibition assay.

##### III. Hemagglutination Inhibition Assay

For the assay, paired RIA vials were prepared for each sample: one serving as Control and the other as Unknown (patient or donor). 50 µL of respective antibody from the 2+ Daughter vial were added to each vial. The Control vial received 50 µL of normal saline, whereas the Unknown vial received 50 µL of preserved saliva. Vials were incubated at room temperature for 10 minutes, followed by the addition of 50 µL of 5% RBC suspension. After gentle mixing, vials were incubated at room temperature for 1 hour and centrifuged at 1,000 rpm for 1 minute prior to visual inspection.

##### IV. Interpretation of Results

Results were interpreted as follows: samples were classified as secretor if no agglutination was observed in any of the three blood group–specific vials (A/B/O). Samples showing agglutination in all three vials were classified as non-secretors, indicating the absence of detectable A, B, or H antigens.

##### V. Result Validation

Positive (non-secretor saliva) and Negative (Bombay saliva) controls from the departmental repository were used in each batch of results.

## Statistical Analysis

Data were analyzed using the Chi-square test to assess the association between secretor status and cancer. A significant association was observed between non-secretor status and cancer (p < 0.0001).

## Results

### Participant Characteristics

A total of 74 female participants were included in the study, comprising 37 cancer patients (23 with breast cancer and 14 with ovarian cancer) and 37 healthy controls. All participants provided saliva samples for secretor status determination. Baseline demographic and clinical characteristics were comparable between the groups.

### Seroprevalence of FUT2-Mediated Secretor Status

The distribution of secretors and non-secretors among healthy controls and cancer patients is summarized in Table 1 and illustrated in Figure 4.

**Table 1.** Distribution of FUT2-mediated secretor and non-secretor status among study participants. Healthy controls showed a predominance of secretors (78%), whereas breast and ovarian cancer patients demonstrated a higher prevalence of non-secretors (82.6% and 78.5%, respectively).

| Group | Total (n) | Secretors n (%) | Non-Secretors n (%) |
| --- | --- | --- | --- |
| Healthy controls | 37 | 29 (78%) | 8 (22%) |
| Breast cancer cases | 23 | 4 (17.4%) | 19 (82.6%) |
| Ovarian cancer cases | 14 | 3 (21.5%) | 11 (78.5%) |

**Figure 4:**
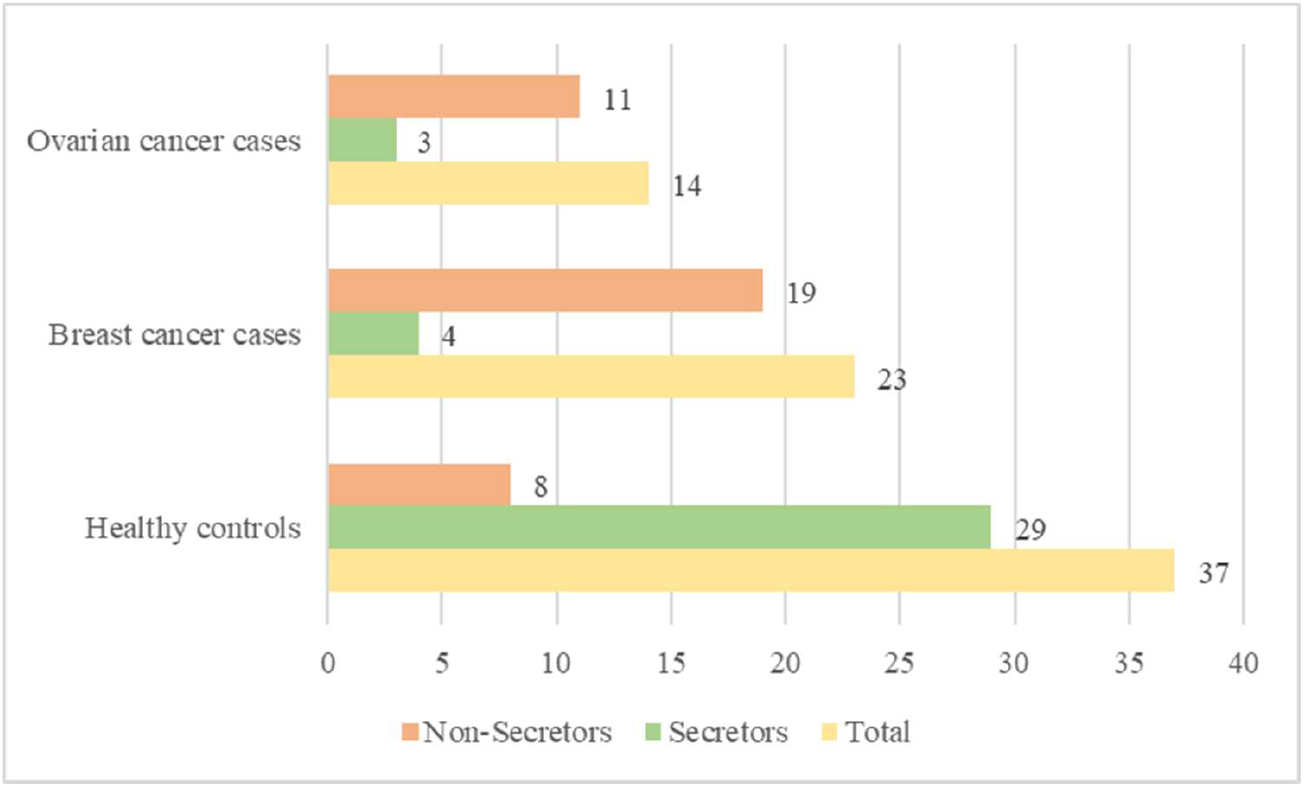
Distribution of FUT2-mediated secretor and non-secretor status among study participants. Bar chart showing the number of secretors and non-secretors among healthy controls, breast cancer cases, and ovarian cancer cases. Non-secretors predominated in both cancer groups (82.6% in breast cancer, 78.5% in ovarian cancer), whereas secretors were more common among healthy controls (78%).

Among the healthy controls, 78% (n = 29) were identified as secretors, while 22% (n = 8) were non-secretors. In contrast, 81% (n = 30) of cancer patients were non-secretors, with only 19% (n = 7) being secretors. Stratification by cancer type revealed that non-secretor prevalence was 82.6% (n = 19) in breast cancer patients and 78.5% (n = 11) in ovarian cancer patients.

### Association Between Non-Secretor Status and Cancer

A significant association was observed between non-secretor status and cancer occurrence (p < 0.0001). Non-secretor status was markedly more prevalent among cancer patients compared with healthy controls, indicating a potential link between FUT2-mediated non-secretor phenotype and susceptibility to breast and ovarian cancers.

## DISCUSSION

In this study, we observed a significantly higher prevalence of non-secretors among breast and ovarian cancer patients, suggesting that absence of α-(1,2)-fucosyltransferase activity encoded by FUT2 may increase susceptibility. This finding aligns with previous evidence from pancreatic and oral cancers, where non-secretor status has consistently been linked to elevated cancer risk.

Oral cancer studies by Rai et al. (2015), Bakhtiari et al. (2019), and Moreno et al. (2009) converge on the finding that non-secretors are more prone to potentially malignant disorders. Complimenting this, Kim et al. (2023) found that secretor status modifies PDAC risk in combination with ABO blood type. These correlations suggest a protective role of mucosal ABH antigen secretion, possibly by maintaining epithelial barrier function or modulating the microbiome. The authors proposed that future research should focus on clarifying the role of FUT2 gene polymorphisms as predictive markers for oral cancer risk, suggesting that non secretors could be more vulnerable due to a lack of wild-type FUT2 gene expression.^13,15,22,23^

However, despite this growing body of evidence across gastrointestinal, pancreatic, and oral cancers, the serological or molecular investigation of secretor status in breast or ovarian malignancies has been limited. This represents an important gap, given that multiple mechanistic studies already implicate FUT2 in carcinogenic pathways. Our findings provide the first serological prevalence data on secretor status in breast and ovarian cancer, revealing a striking disparity with over 70–80% non-secretors among patients.

FUT2 overexpression has been observed in autophagy and inhibition of apoptosis through regulation of AMPK, ULK1, PI3K III, DRAM1, and JNK pathways, thereby promoting tumor cell survival. Complimenting this, Deng et al. (2018) showed that FUT2 facilitates epithelial– mesenchymal transition (EMT) by fucosylating TβRII and activating the TGF-β/Smad pathway, contributing to metastasis. Together, these studies highlight FUT2’s role in supporting both survival and invasion, but important questions remain regarding how these processes are coordinated.^24,25^

Addressing some of these gaps, Dong et al. (2024) demonstrated that FUT2 reprograms lipid metabolism by activating YAP/TAZ signaling and stabilizing mSREBP-1, thereby driving fatty acid synthesis to support colorectal cancer metastasis.^26^

Beyond these molecular functions, tissue-specific studies highlight FUT2’s diverse clinical associations. In head and neck squamous cell carcinoma (HNSCC), Montesino et al. found high FUT2 expression correlates with better survival, through preservation of LeY antigens on immune-regulatory proteins.^27^ In a 2017 study, it was identified that the rs1047781 T allele correlates with more advanced clinical stage, larger tumor size, and higher AST/ALT ratio in hepatocellular carcinoma patients, while Hung et al. (2022) identified FUT2 as a key enzyme contributing to the overexpression of the tumor-associated glycan Globo H.^18,28^

Non-secretors are unable to secrete the A/B/H antigens in their bodily secretions and that can lead up to gut dysbiosis when compared to a secretor. These antigens can influence the microbial diversity and abundance by acting as substrates for various commensal microbes such as *Bifidobacteria* and *Lactobacilli*. A reduction or absence in beneficial species is known to contribute to the progression of colorectal and other cancers through mechanisms such as chronic inflammation, microbial metabolite production, and genotoxic effects.^29,30^

The ease of non-invasive, saliva-based testing also underscores the translational potential of secretor status as a marker for risk stratification making it feasible for population-level studies, including in low-resource settings.^31^

In the present pilot study, a larger sample size with more diverse population along with the analysis of the molecular polymorphisms of the FUT2 gene could have been more effective to provide better insights by which salivary non-secretor status might be a potential risk factor for breast and ovarian cancers. This was a limitation of our present study.

## Acknowledgement

The authors acknowledge the cooperation of all the Junior resident doctors, Senior resident doctors, Medical officer - in – charge, faculties (Dr. B. Talukder, Dr. C. Maity) and Ms. Shreya Jha of Department of Immunohematology and Blood Transfusion, Medical College Kolkata.

## Data Availability

The data generated and analyzed during this study are not publicly available due to privacy considerations. Researchers may request access to the de-identified dataset from the corresponding author, subject to institutional review and ethical approval.

